# Immediate and Near Future Prediction of COVID-19 Patients in the U.S. Population Aged 65+ With the Prior Medical Conditions of Hypertension, Cardiovascular and Lung Diseases: Methods, Models and Acute Care Estimates

**DOI:** 10.1101/2020.04.12.20062166

**Authors:** Arni S.R. Srinivasa Rao, Douglas D. Miller, Adam E. Berman, David C. Hess, Steven G. Krantz

## Abstract

**Importance:** Given the rapid rise of COVID-19 cases in the U.S. during March 2020 there has been a severe burden on the health care systems and care providers in the country. The impact of the virus so far was higher on the population aged 65+. Hospitalizations were higher among those with underlying medical conditions, namely, hypertension, cardiovascular and lung diseases. Hence, to have an idea of the number of new COVID-19 infections among these high-risk populations that could occur in the short-term could assist promptly to the country’s health care system for immediate health care planning. These estimates may aid us in better understanding the potential volumes of patients requiring inpatient care.

**Objective:** To provide immediate and short-term model-based predictions of COVID-19 patients in the U.S. population aged 65+ during April-June, 2020, those with the prior medical conditions of hypertension, cardiovascular and lung diseases.

**Design, Setting, and Participants:** We developed age-structured dynamic mathematical combined with wavelet analysis to understand the number of new cases that may emerge in the U.S. population aged 65+. We have estimated the number of people aged 65+ who might have three underlying conditions mentioned and a possible number of hospitalizations among them due to COVID-19 if they get infected. We have used publicly available data sources for developing our framework and estimates.

**Results:** We estimate that there are 13 million individuals aged 65+ who have one or a combination of three major prior medical conditions in the U.S. who need to be protected against COVID-19 to reduce a large number of hospitalizations and associated deaths. Hospitalizations of patients both with and without ICU-admissions with more prevalent underlying conditions could range between 31,633 (20,310 non-ICU hospitalizations and 11,323 ICU-admissions) to 94,666 (60,779 non-ICU hospitalizations and 33,866 ICU-admissions) cases during the same period. Under a rapid spread of the virus environment, these hospitalizations could be beyond 430,000 within the above three-month period.

**Conclusions and Relevance:** COVID-19 continues to dramatically and adversely affect the lives of people aged 65+ in the U.S. During the next three months which could result in thousands of hospitalizations if precautions against the virus spread are not implemented and adhered to.

## Introduction

The novel coronavirus, SARS-Cov-2, is the agent responsible for COVID-19, and has caused more than 10,000 hospitalizations within a short period since the first case was detected in the U.S. on January 20, 2020, in a returned traveler from Wuhan, China. On February 26th, the CDC (Centers for Disease Control and Prevention) reported the first case from a person who had no travel history or no known contact with an infected individual [1]. As of April 10, 2020, the number of reported cases in the U.S. is 505,015 with more than 18,771 deaths and 29,000 recovered cases. Globally, there have been more than 1.7 million people infected as of April 10, 2020, with 104,000 deaths.

CDC derived hospitalization data on COVID-19 patients in the U.S. demonstrates that 89.3% of affected individuals had underlying conditions [2]. The most common of these underlying conditions were hypertension (49.7%), obesity (48.3%), chronic lung disease (34.6%), diabetes mellitus (28.3%), and cardiovascular disease (27.8%). A MMWR report released by CDC on April 3, 2020 [3] indicates that among U.S. adults aged 19+ who were hospitalized due to COVID-19 through March 28, 2020, almost 60% were of age 65+ who were reported to have one or more underlying medical condition. Some of these hospitalizations resulted in admission into an Intensive Care Unit (ICU) setting. In the April 8^th^ MMWR report [2], it was mentioned that among the U.S. COVID-19 cases aged 65+, the most prevalent prior conditions were hypertension (72.6%), cardiovascular diseases (50.8%), diabetes (45%) and lung diseases (38.7%). Eighty percent of the U.S. COVID-19 related deaths reported through March 16^th^ were aged 65+ [4]. Several other U.S. and international research studies have indicated COVID-19 has a strong association with older adults requiring hospitalization if they also possessed any prior underlying medical condition [5-11].

Considering the importance of infections among older adults, especially those aged 65+ who exhibit a higher probability of hospitalization if becoming infected with COVID-19, we undertook our study to predict the possible number of people in this age group who might become infected during next three months (April-June, 2020) in the U.S. We aim to provide these predictions with a breakdown of the possible number of patients who also exhibit the underlying comorbidities of hypertension, cardiovascular diseases, and lung diseases. We developed mathematical methods and dynamic models to predict the number of persons who might be COVID-19 infected between April 7 and June 30, 2020. Our previous modeling work on COVID-19 has contributed important finding on under-reporting in the U.S. and in other countries [12-13], policy frameworks [14]. Our COVID-19 research has also contributed to the development of deeper mathematical methods for obtaining true epidemic growth through wavelet functions [15] and artificial intelligence modeling frameworks for identification of COVID-19 [16]. Details on the methods developed, models, and the data are provided in the Appendix. We have provided three sets of model-based predictions, namely, lower, medium and higher, with corresponding numbers of underlying medical conditions in the U.S. among aged 65+.

## Key findings

We predict that there will be at least 34,000 to 104,000 more infections among the U.S. 65+ year old population during April 7 and June 30, 2020. This range could extend to 470,000 more infections due to higher transmission rates if insufficient precautions against the spread of the virus are implemented (**Figure 1**).

**Figure 1.**
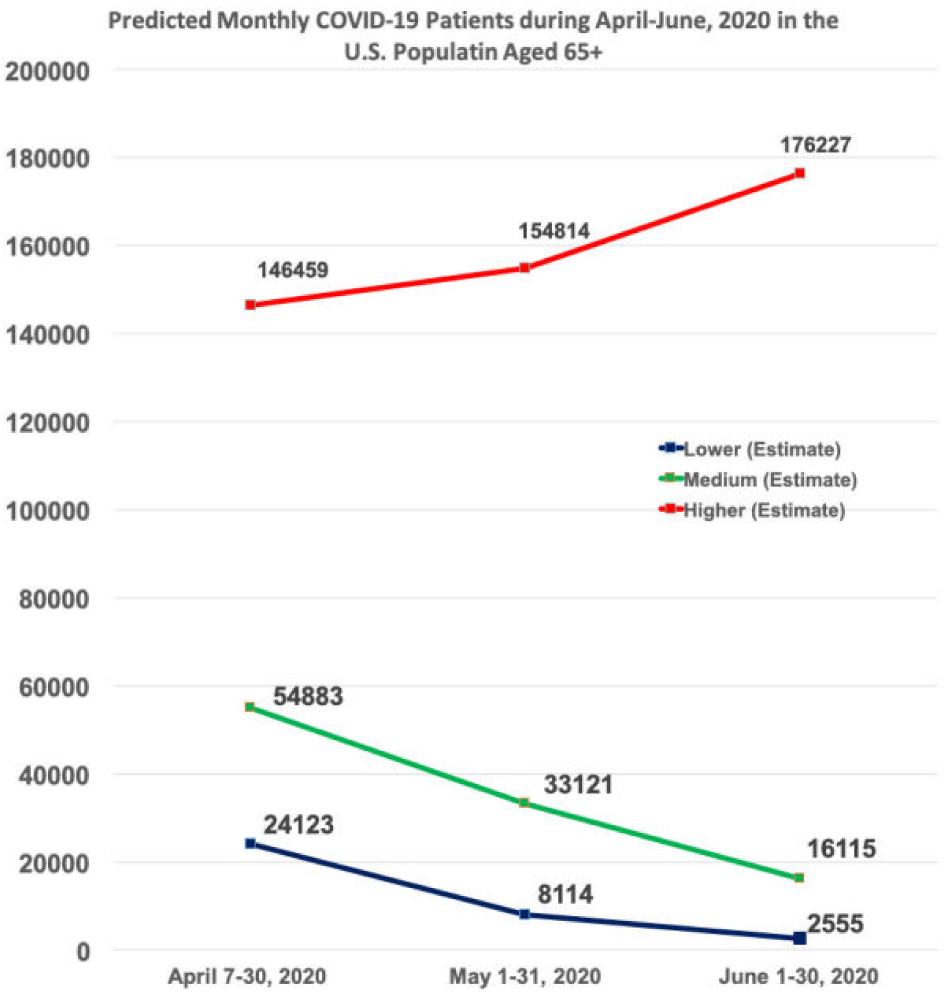
Predicted Monthly new COVID-19 Patients in the U.S. Population aged 65+ during April–June, 2020.

Our predictions on hospitalizations during this period of people aged 65+ range between 31,000 to 94,000. Under a relaxed spread environment, these hospitalizations could be beyond 430,000 within the above three-month period. See the breakdown of these hospitalization numbers with and without ICU-admissions in **Figure 2** and **Table 2**.

**Table 2.**
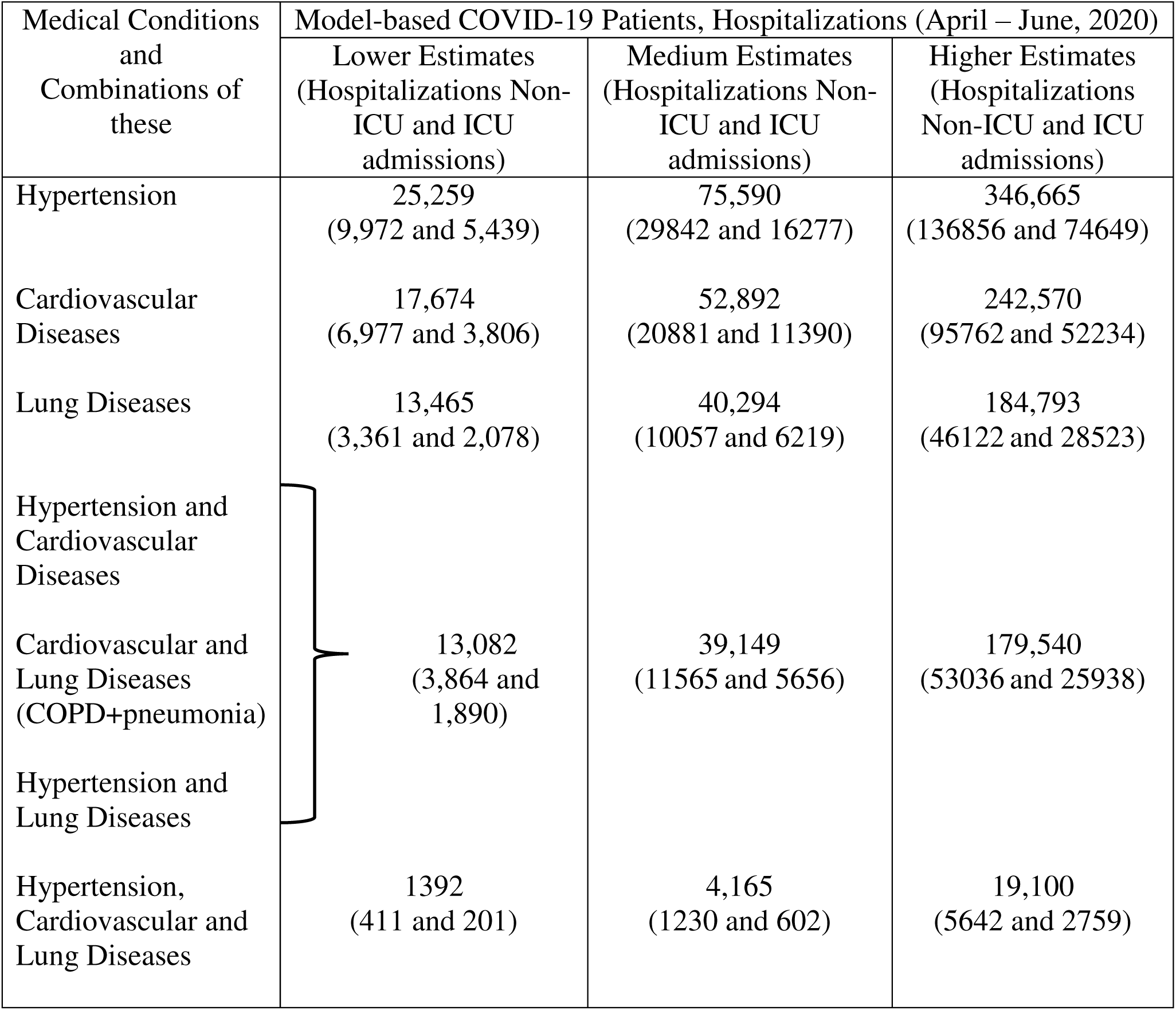
Predicted COVID-19 Patients, Hospitalizations with and without ICU admissions among the U.S. Population aged 65+ during April 7 – June 30, 2020 who will have Prior Medical Condition or Combinations of Medical Conditions.

**Figure 2.**
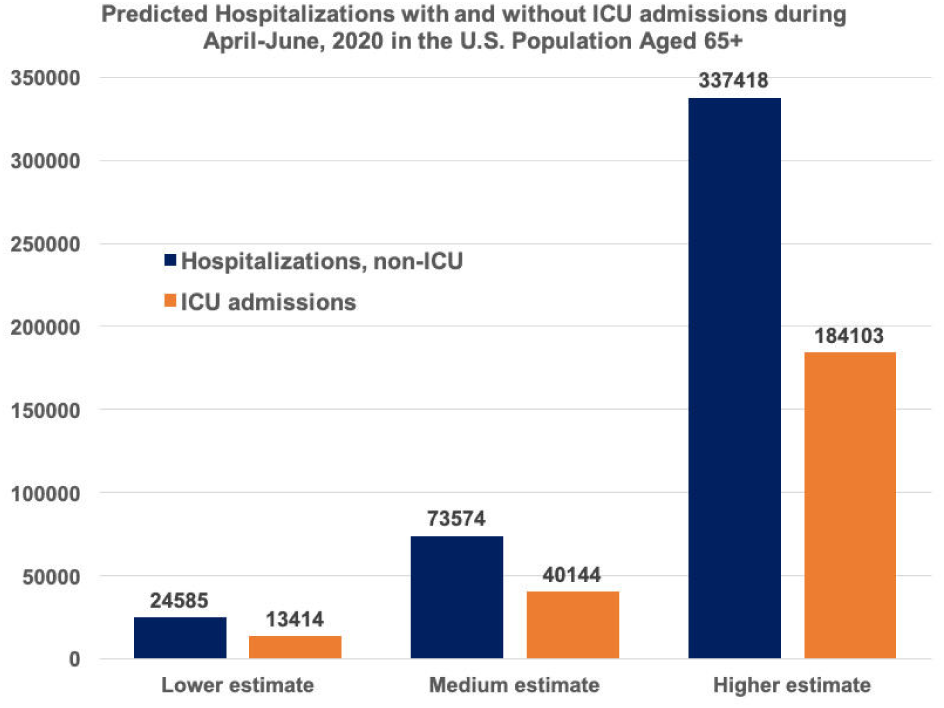
Predicted hospitalizations with and without ICU admissions during April-June, 2020 among the U.S. population aged 65+.

Our predicted hospitalizations with a breakdown of underlying conditions indicate that there will be at least 25,259 (9,972 non-ICU and 5,439 ICU) hospitalizations of hypertension-related, 17,674 (6,977 non-ICU and 3,806 ICU) of cardiovascular disease-related, and 13,465 (3,361 non-ICU and 2,078 ICU) cases of lung diseases related hospitalizations during April 7 and June 30, 2020. These hospitalization numbers for medium-range predictions are 75,000, 53,000 and 40,000 for hypertension, cardiovascular and lung disease-related, respectively. **Table 2** provides further details of these estimates, including the composition of a potential worst-case scenario.

Our predicted hospitalizations with a breakdown of underlying conditions indicate that there will be at least 15,400 cases of hypertension-related, 10,700 cases of cardiovascular disease-related, and 5,400 cases of lung diseases related hospitalizations during April 7 and June 30, 2020. These hospitalization numbers for medium-range predictions are 46,000, 32,000 and 16,000 for hypertension, cardiovascular and lung disease-related, respectively. **Table 2** provides further details of these estimates, including the composition of a potential worst-case scenario.

We have demonstrated the magnitude of the difference between various underlying conditions using (Yves) Meyer’s wavelets, which are very flexible to capture within a broader spectrum of values. Meyer wavelets are a Fourier transformation of a scaling function, useful for multi-fault classification based on signal decomposition levels using wavelet analysis and support vector machine learning (**Figure 3**).

**Figure 3.**
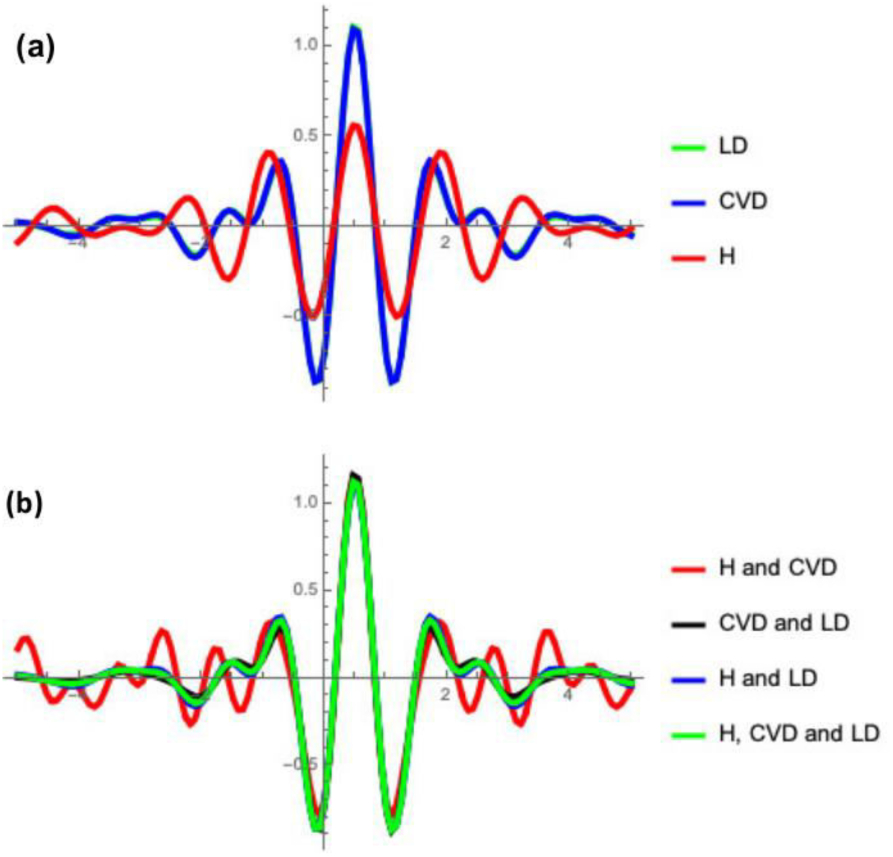
Meyer’s wavelets of number of predicted prior conditions among COVID-19 patients in the U.S. population aged 65+ during April – June, 2020.

**Figure 4.**
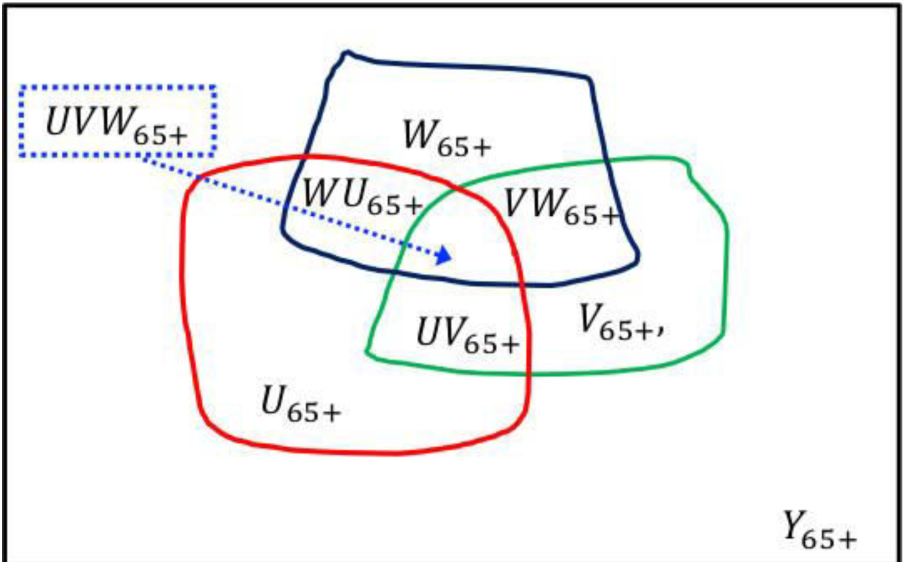
COVID-19 patients (*Y*_65+_) who are aged 65 and above and subsets of these patients with one or more of the prior medical conditions. Here *U*_65+_ *V*_65+_ and *W*_65+_ are the subsets of COVID-19 patients people (*Y*_65+_) with prior medical conditions, hypertension, cardiovascular disease, and lung diseases as of the year 2020. *UV*_65+_ *VW*_65+_ and *WU*_65+_ are the corresponding subsets of patients with pairs of medical conditions (hypertension and cardiovascular diseases), (cardiovascular and lung diseases), and (lung diseases and hypertension), respectively. *UVW*_65+_ are the patients who have all three prior medical conditions within overall COVID-19 infected aged 65+, i.e. (*Y*_65+_).

**Figure 5.**
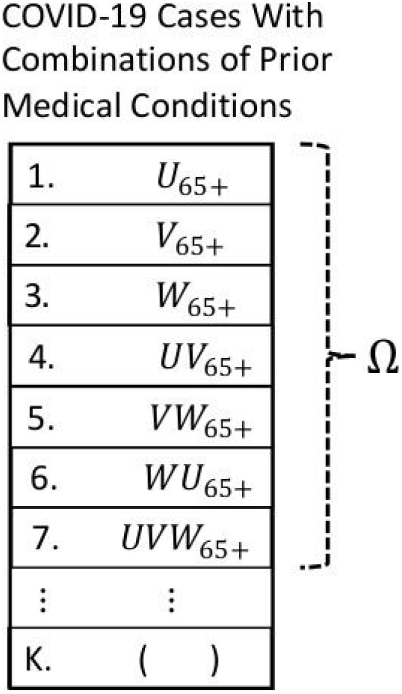
COVID-19 cases with a list of all possible Individual and combinations of prior medical conditions. Suppose there are *K* items in that list. We have in this study considered |Ω| number of such combinations with |Ω| < *K*. We could extend the study with a larger |Ω| such that |Ω| → *K* if we have the evidence of such data on prior medical conditions.

## Conclusions

COVID-19 continues to dramatically and adversely affect the lives of people aged 65+ in the U.S. During the next three months which could result in thousands of hospitalizations if precautions against the virus spread are not implemented and adhered to. When evaluating death rates attributable to COVID-19, it is important to separate hospitalizations caused solely by COVID-19 from hospitalizations caused by other illnesses and underlying conditions. This information is important for epidemiological and public health planning purposes, and also for a full understanding of the spread of the disease. Our predictions will be useful in providing state level predictions so that more in-depth prevention measures may be developed and implemented.

## Data Availability

Publicly available data is used.

## Appendix: Methods, Models and the Data

Let *U, V* and *W* be the three sets representing the number of individuals in the USA who are having prior medical conditions: hypertension, cardiovascular diseases, and lung diseases as of the year 2020. Let *U*_65+_ *V*_65+_ and *W*_65+_ be the corresponding sets of people who are aged 65 or above. Let *UV*_65+_ *VW*_65+_ and *WU*_65+_ be the corresponding sets of people who are aged 65 or above and have pairs of medical conditions (*U, V*), (*V, W*) and (*W, U*), respectively. There will be a smaller class of people who are 65 + and having all three medical conditions *U, V* and *W*, which we denote by *UVW*_65+_. All these combinations of sets of prior medical conditions are defined below:

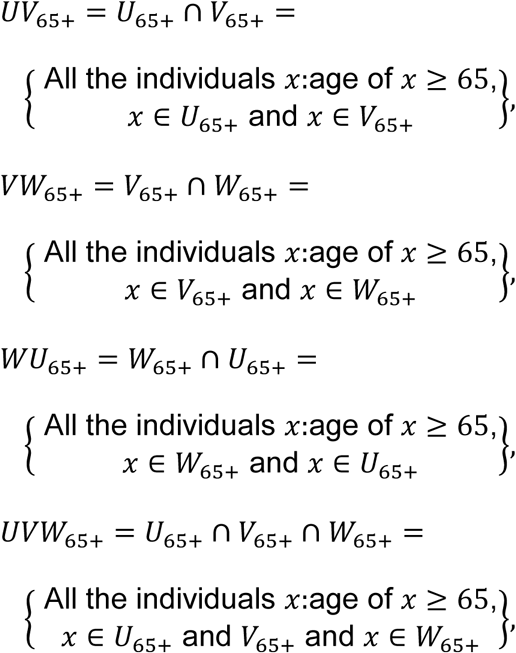

Let Ω be the set of all the sets of prior medical conditions, given by, Ω

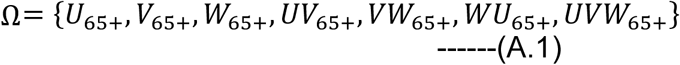

Once we have these sets of people with prior medical conditions, we will obtain the number of COVID-19 infected in the U.S. at time *t*, say, *Y*_65+_ (*t*), by solving numerically coupled differential equations in (A.2),

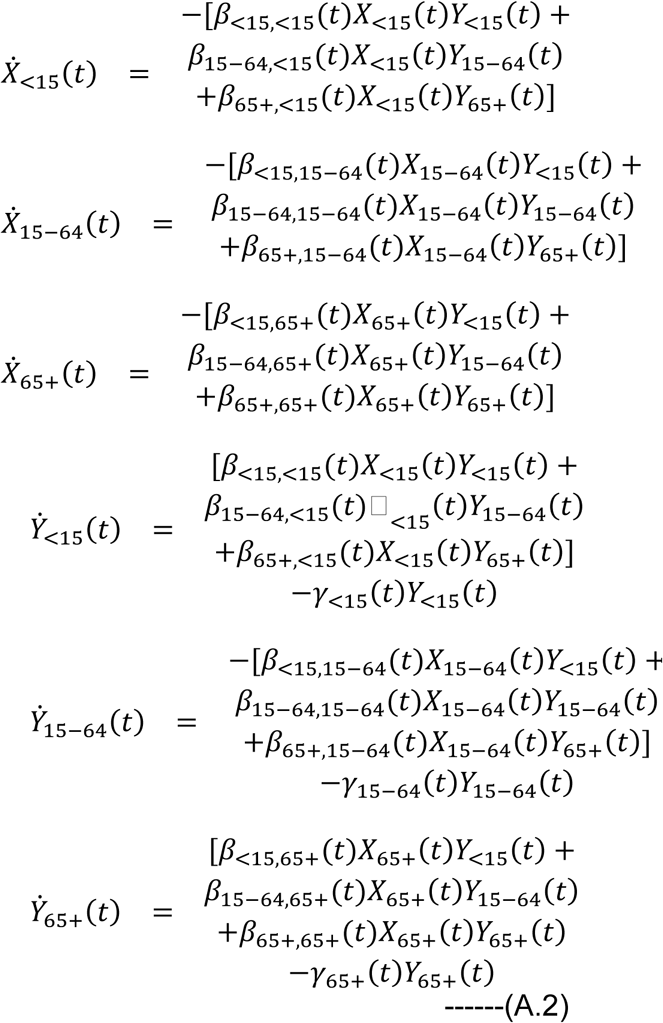

Here *X*_<15_, *X*_15−64_, *and X*_65+_ represent susceptible people in the age-group<15, 15-64, 65+, respectively at time *t. Y*_<15_ (*t*), *Y*_18−64_ (*t*) and *Y*_65+_ (*t*) represent those who are COVID-19 infected in the age-groups < *15, 15* − *64*, and 65+, respectively at time *t*. Here *β*_*ij*_ (*t*) represent the average transmission rate from an infected in the age group *j* to an susceptible in the age group *i*. for *i, j = 15, 15* − *64*, and 65+, respectively at time *t*. The recovery rate parameter is *γ*_*i*_ (*t*) for the age-groups <15, 15-64 and 65+ at the time *t*. Dynamic models without the age structure populations were used by us for the COVID-19 [12-13]. Age-structure deterministic models were also built for understanding other epidemics, such as, HIV/AIDS, bird flu, Hepatitis, other diseases and in population dynamics, see for example in [17-30]. The set of all transmission rate parameters, say, *β*_0_, can be visualized as

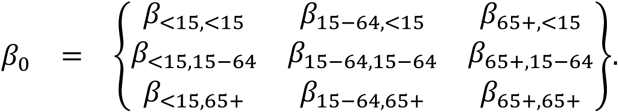

We have assumed *β*_*ij*_ *= β*_*ji*_ for *i* ≠ *j* because the interacting populations will have same people and we do not have much information on the age-structure of the transmission rates. This will reduce □_0_ into a new set β given below:

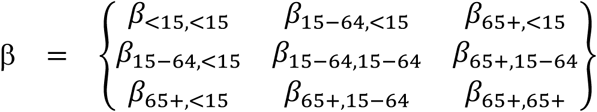

Based on the transmission values considered in our earlier articles on COVID-19 and further calibrating for the U.S. reported trend observed and model-based predictions for the periods March 1-14, 2020 and March 15-April 6, 2020, the β was considered for the medium range. Then the lower values and higher values were reduced to a fraction values of the medium range and the higher values increased to the same fraction above the medium range.

We will predict the number of individuals in each of the sets in Ω. Before we do this, for each element *A* in Ω, we assume

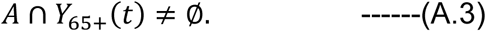

That is, there are at least one or more individuals in each of the sets *A* in Ω who is COVID-19 infected. Even if we violate the assumption by having one or more sets *B ≠ A* in Ω such that

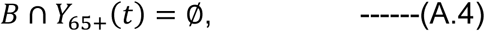

then we provide model-based estimates for those sets which satisfy (A.3).

From the U.S. COVID-19 data, we will obtain the fraction of cases who will have prior medical conditions for each element of Ω. These will form the initial values of each of the sets. We will also form monthly averages of these fractions. We simulate *γ*_65+_ (*t*) values over the period April 7-June 30, 2020 and multiply the proportions of each of the sets of Ω after the time period of data collection is adjusted for monthly rates of infections within prior conditions. Suppose that *f*_*i*_ (*A*) is the fraction of the set *A* of those who were infected with COVID-19 within the *i*^*th*^ month, then, by computing

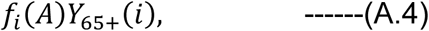

for *i =* 1,2,3, we will obtain the monthly number of individuals who will likely acquire with prior medical condition *A*. Here *Y*_65+_ (*i*) is the number of COVID-19 cases for *the i*^*th*^ *month*. Note that

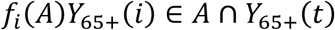

for each *A* ∈ Ω. Hence, 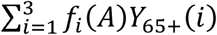 represents the number of individuals within each prior medical conditions who need to be protected from being infected. These will assist in preventing death rates among such individuals.

From the magnitude of possible cases predicted for each of the sets in Ω, the Meyer wavelets were constructed. The Meyer wavelets *ψ*(*ω*) can be treated as infinitely differentiable functions within a certain domain. See details in [12-13,15]. The Meyer wavelets *ψ*(*ω*) and their accompanying function *φ* are given below

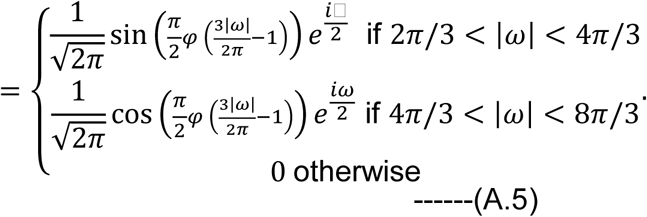

Here *φ* (*x*) = 0 for *x* < 0, and

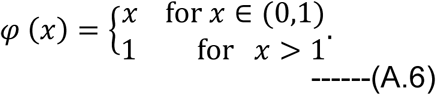

We plotted Meyer wavelets for the difference of the magnitude of the COVID-19 for the three-month period mentioned. These will help us in seeing the comparative dynamics of possible impact due to COVID-19 in terms of underlying conditions and hospitalizations (as shown in Figure 6). Wavelet theory is an improvement over traditional Fourier series expansions *f*(*x*) defined either on [*0*,2*Π*) or on [0, *Π*) using *sin* and *cos* functions, where

**Figure 6.**
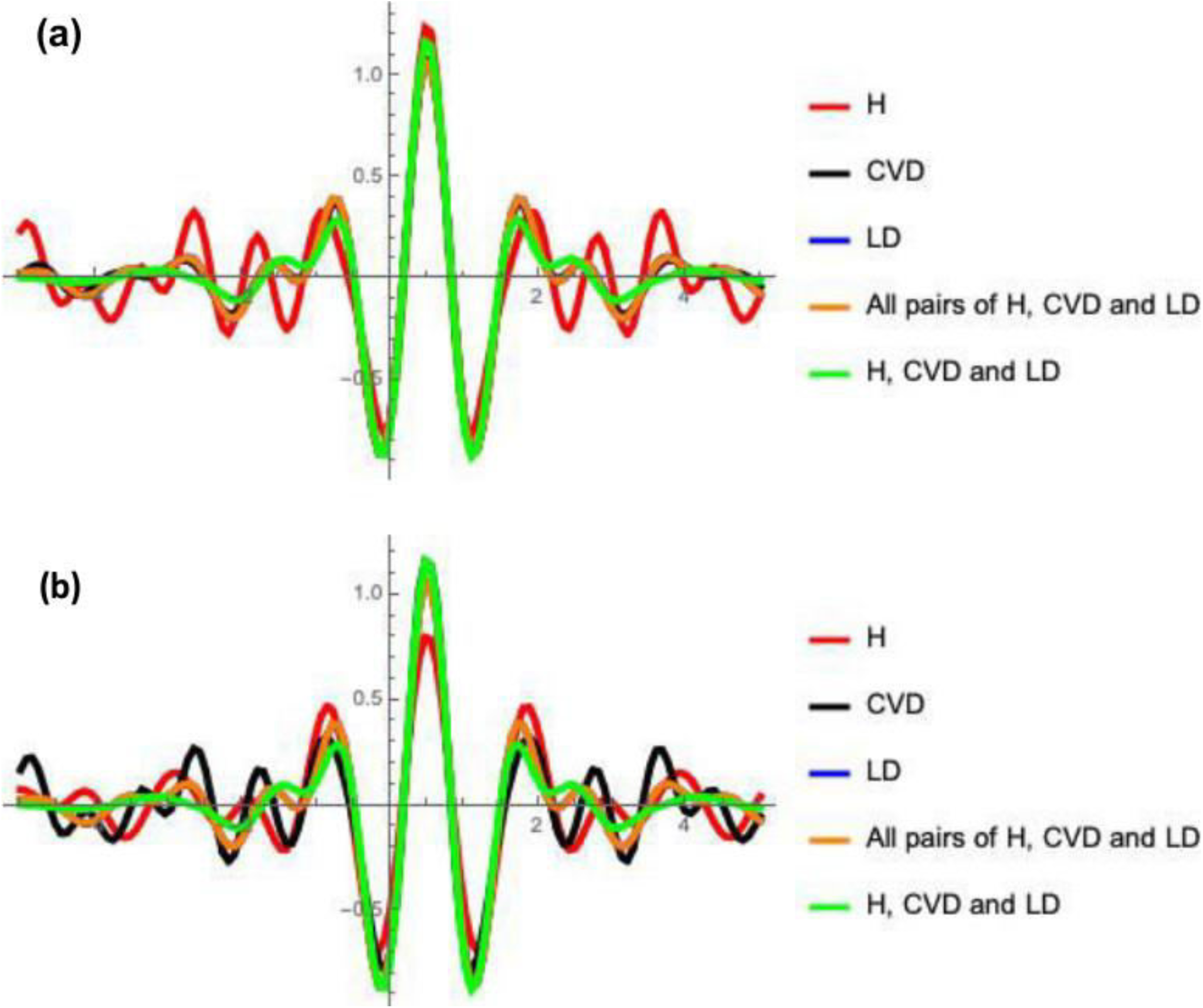
Meyer’s wavelets of COVID-19 estimates for the period April-June, 2020 and Hospitalizations (a) COVID-19 estimates (b) Hospitalizations

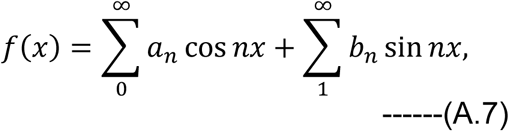

where *a*_n_ and *b*_n_ are coefficients of the series. Wevelets are shown to have immense help in several fields, including in epidemiology [15]. Further details and applications on wavelets can be seen elsewhere, for example in [31-35].

The entire methodology described can be repeated by applying other age group data as well as other prior medical conditions data.

## Data

### Heart-related hospitalizations

The heart-related hospitalizations were estimated for 2016 data published in 2018 [36]. We have summed up the numbers for men and women in the age groups 65-74 and 75+ to obtain 65+ numbers, which is 1,432,000. Then we have assumed annual growth of these numbers at a rate of 0.7% to obtain the value of 1,472,519 for the year 2020. There were other studies that estimate burden of hospitalizations due to heart-related complications [37-44].

### Hypertension

According to Older American’s profile of 2017 published by ACL [44] (Administration of Community Living) within the department of health and human services, 58% of the US population aged 65+ are having hypertension. We have applied this percentage to the US population who are aged 65+ for the year 2020 [45] to obtain the number of individuals who are aged 65+ in the U.S. who have hypertension. We have applied hypertension percentage estimated for the year 2017 on the 2020 population (as there was no published estimate of hypertension percentage among persons aged 65+ for the year 2020). According to [46], the U.S. population estimate for the year 2020 is 331 millions of which 14.3% are aged 65+ [45]. This gives us the size of the U.S. population 65+ are 47.33 million and 58% of them, that is, 27.45 million with hypertension.

According to the data by the CDC, there are 33.5% of the persons aged 20+ in the U.S. who are taking medicine for hypertension during 2015-2016 [47]. Other studies indicated rise of hypertension among adults in the U.S. and in other countries [48-57].

### Lung related illnesses

We have divided these broadly into two categories COPD (Chronic obstructive pulmonary disease) and pneumonia. According to [58], the global burden of hospitalizations due to COPD for aged 65+ is 14.2% and we do not have such statistics for the U.S. Applying this percentage of the US population aged 65+ gives us that there will be 6,721,286 people who are at risk of COPD in the U.S. in 2020. Among the COPD patients who are aged 45+, 51% admitted to the hospitals from an index ED visit [59]. Approximately 28.6% of the 45+ will be in the age group 65+ (as 14.3% of the total are in the age-group 65+). These fractions gives us that potential hospital visits due to pneumonia for the year 2020 will be an estimated 980,367.

According to the ambulatory and hospital care statistics survey in the US [60], there were 1.3 million visits to various emergency departments. We do not have information how many of these visits are by different individuals. An annual growth of 0.7% of those numbers as per the population growth in the country gives us an estimate of 1.59 millions for 2020. If we assume population proportion of aged 65+ to these numbers, then the anticipated hospital visits will be approximately 227,735 for the year 2020.

Overall lung-infections-related hospital visits will be 1,208,103.

### Comorbidity of Hypertension and Cardiovascular Diseases

Individuals with hypertension have several comorbid conditions with heart related diseases in the U.S. and other countries [61-66]. For example, medical expenditure survey data of 2011-2014 [67] indicates, individuals with hypertension have the following comorbid conditions: coronary heart disease (16.7%), heart rhythm disorders (6.0%), congestive heart failure (2.1%). This gives us 6,808,379 individuals who have comorbidity of hypertension and cardiovascular diseases in the U.S. for the year 2020.

### Comorbidity of Cardiovascular Diseases and Lung diseases

Several studies have shown COPD with prevalence of comorbidity with cardiovascular diseases [67-71]. We consider the comorbidity prevalence 18% reported from the study [70] because the sample size they considered was one of the largest. The estimated comorbidity of COPD with cardiovascular diseases is 176,466 for the year 2020.

Other studies indicate pneumonia could worsen cardiac conditions [72-82]. In the study [83] it was found that 12% of the hospitalized with pneumonia had cardiovascular diseases. By applying this percentage of the CVD patients who are aged 65+ gives us 176,703 patients are at risk of hospitalizations in the year 2020.

### Comorbidity of Hypertension and Lung diseases

The comorbidity due to COPD is one of the important and well-studied medical conditions [84-95]. The study [87] reports 28% of the COPD have comorbid condition of hypertension. A study by [85] considered 20,296 COPD patients to study comorbid conditions with diabetes, hypertension and cardiovascular diseases. Out of 437, 1680, 1168, 1670 patients aged 65-71, 72-75, 76-79, >80, the hypertension was recorded in 49.9%, 51.3%, 58%, 62.4%, respectively. A simple weighted average gives us 56.5% of the COPD patients have hypertension as a comorbid condition. Applying these percentages on the COPD estimates of 2020 obtained above gives us 553,907 patients will have comorbid conditions of COPD and hypertension.

Hypertension has been found to be associated with pneumonia related hospitalizations in the US and several other countries [96-99]. We used 2.1% of the hypertensive individuals will have pneumonia during the year, which comes to 576,516 individuals.

### Multiple morbidity among older patients

Several studies are available on multiple morbidlity data on older adults [100-104]. A study by Skinner et al [101] finds 30.1% of the elder population aged 65+ have 4 or 5 morbidities require hospitalization. We have applied these findings on the populations of hypertension and lung diseases combined and cardiovascular diseases and obtained an estimate of 783,486 for the year 2020.

### Age-distribution of reported cases and model-based numbers in the U.S

According to CDC’s MMWR released on April 2, 2020 [105], out of the total COVID-19 cases reported until April 2, 2020 and whose age is known, there are 1.7% aged 18 or less. As of April 6, 2020, there are 367,004 COVID-19 cases reported in the U.S. Since the estimate for infected in the 0-15 age is not available, we use applied above percentage on the modeling based numbers obtained for the period. According to the same MMWR report, 76% of the infected persons are aged 18-64, and the remainder 22.3% are aged 65+. Our earlier model-based estimate as of April 6 had 561,005 COVID-19 cases in the age group 0+ and out of which 146,840 are in the age group 65+. These model-based numbers include reported and not reported. Assuming detected cases and recovered cases are under quarantine or taking precautions not to spread the disease to others, we have 561,005×1.7%-367,004×1.7% = 3298 persons aged 0-14 who will be contributing to the spread after April 6, if not taking any precautions. Similarly, 561,005×76%-367,004×76%=147,441 persons aged 15-64 who will be contributing to the spread and 561,005×22.3%-367,004×22.3%=43,262 persons aged 65+ will be contributing to the spread after April 6, 2020 if proper care is not taken. There are 51,374,510 individuals aged 0-14, 179,948,150 individuals aged 15-64 and 43,407,340 individuals aged 65+. Subtracting those individuals who were already detected from these age groups of susceptible individuals, we will have 51,368,270, 179,669,227 and 43,325,498 numbers of individuals remaining in the susceptible pool.

### Break-up of underlying conditions among predicted COVID-19 cases

Once we have obtained three sets of model-based COVID-19 estimates (lower, medium, and higher) for the period April 7 – June 30, 2020, we have applied the percentages of three underlying conditions and multiple conditions available from [2,3]. We have adopted 4% of the COVID-19 will have all three underlying conditions. Since a detailed break-up of pairs of comorbidity percentages due to hypertension, cardiovascular and lung diseases was not available in the studies [2,3], we have adopted applied 37.8% for all three pairs combined. In the study [3], it was reported that 37.6% of the patients reported having one or more conditions. In general, it is observed in various studies that 4% of the population will have multiple morbidities of 3 or above conditions (Table 2).

### Meyer wavelets

From the predicted morbidities and comorbidities for the period April-June, 2020, the number of COVID-19 numbers predicted with prior medical conditions and predicted hospitalizations in Table 1 and Table 2, we have computed relative magnitudes of each medical condition with the total numbers, and constructed Meyer wavelets (Figure 3 and Figure 6).

**Table 1.**
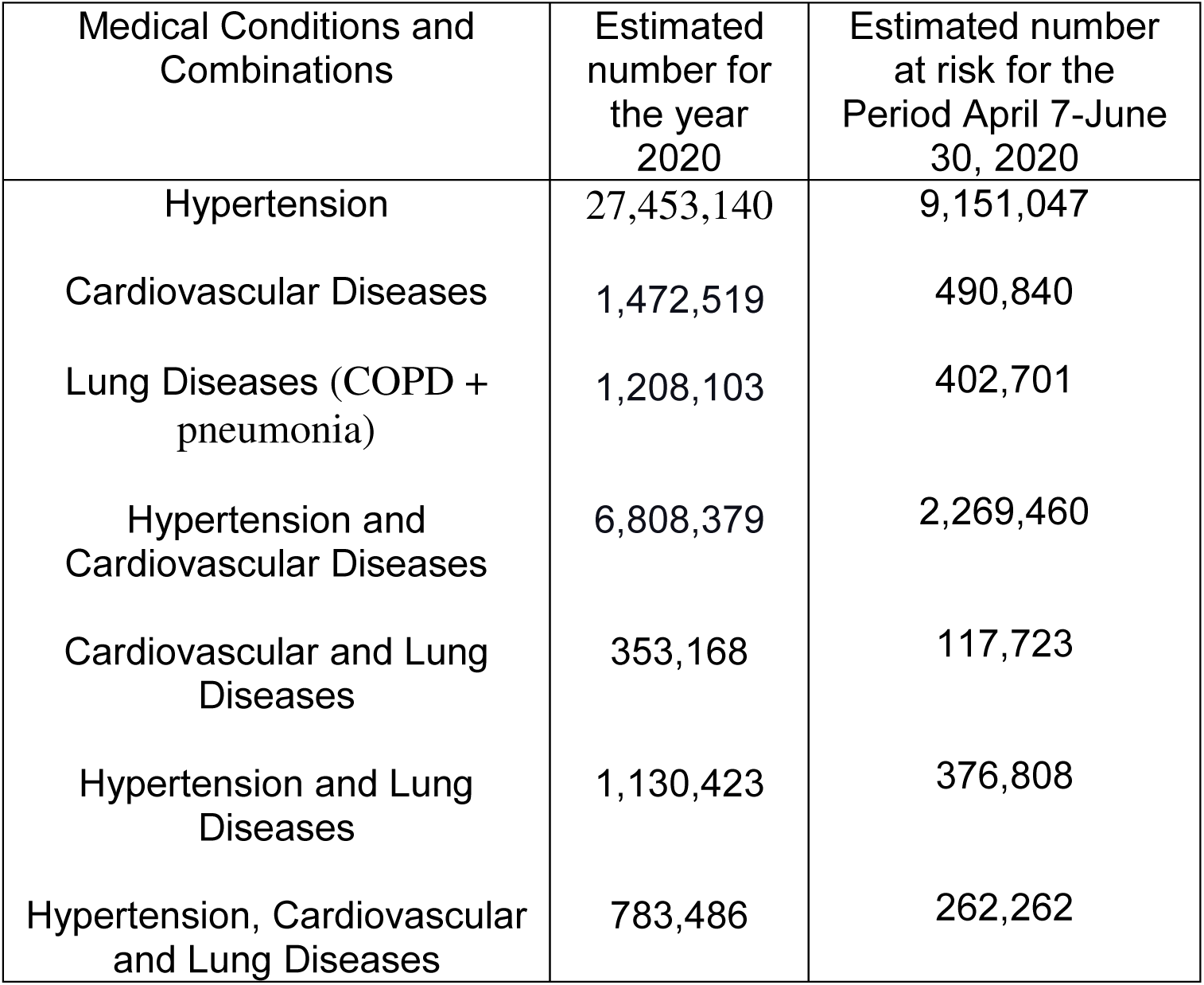
Estimated Prior Medical Condition or Combinations of Medical Conditions for Individuals aged 65+ in the U.S. in 2020 and during April 7 – June 30, 2020 who have risk of hospitalized if infected with COVID-19.

## Acknowledgements

It is a pleasure to thank Randi Diane Ruden for suggesting the topic of this paper.

## Authors contributions

All the authors contributed in writing. ASRS Rao designed the study, developed the methods, models, collected data, performed analysis, computing, creating Figures and Tables, wrote the first draft. D. Miller, A. Berman and D. Hess have contributed in editing, commenting and discussions. SG Krantz participated in the design with ASRS Rao and contributed in editing the draft and providing technical inputs on the methods and models. All the authors approved the final manuscript.

